# Body Mass Index and Incident Tuberculosis in Close Tuberculosis Contacts

**DOI:** 10.1101/2025.04.15.25325878

**Authors:** María B. Arriaga, Gustavo Amorim, Marina C. Figueiredo, Cody Staats, Afrânio L. Kritski, Marcelo Cordeiro-Santo, Valeria C. Rolla, Peter F. Rebeiro, Bruno B. Andrade, Timothy R. Sterling, the RePORT-Brazil consortium

## Abstract

**Background:** Approximately 95% of persons infected with *M. tuberculosis* do not progress to tuberculosis (TB) disease. Identifying key determinants of TB progression could focus prevention efforts.

**Methods:** Contacts of pulmonary TB cases were enrolled in a prospective multi-center cohort study (RePORT-Brazil) from 2015-2019 and followed for 24 months. Dimension reduction techniques included empirical review and LASSO regression, using clinical and laboratory information at baseline, to determine factors for inclusion in prediction models. Models were created for: 1) all contacts, 2) contacts IGRA-positive at baseline, and 3) IGRA-positive contacts who did not receive TB preventive therapy (TPT; <30 days isoniazid). Internal validation was performed using bootstrapping.

**Results:** Among 1846 contacts of 619 TB index cases, 25 (1.4%) progressed to TB. No TPT was a risk factor for progression to TB among all contacts [mixed-effects adjusted hazard ratio (aHR): 11.79 (95% confidence interval (CI): 1.55-89.77). Internal validation of the model with all contacts estimated an area under the ROC curve: 0.85 [95%CI: 0.78-0.91]. Body mass index (BMI) was inversely associated with increased risk of progressing to active TB among IGRA-positive contacts who did not receive TPT (aHR): 0.87 (95%CI:0.78-0.98); IGRA-positive contacts with BMI<25 kg/m^2^ had 4.14-fold (95%CI:1.17-14.67) higher risk of progression to TB than IGRA-positive contacts with BMI ≥25 kg/m^2^; TB risk was 8.4% vs. 2.1%, respectively.

**Conclusions:** BMI<25 kg/m^2^, an easily obtained biomarker, identified IGRA-positive close TB contacts at high risk of progressing to TB disease. TPT should be targeted to this high-risk group to maximize TB prevention.

## Introduction

Although tuberculosis (TB) can be prevented, treated, and cured, it continues to be a major global public health problem [1]. The World Health Organization (WHO) estimated that in 2023 there were 10.8 million cases of incident TB and 1.25 million deaths in persons diagnosed with TB [1]

*Mycobacterium tuberculosis* (Mtb) is transmitted via the respiratory route, from persons with TB to close contacts who share the same space. Exposure to TB can lead to Mtb infection (TBI) [2]. Approximately one-quarter of the global population is estimated to have TBI [3]. The lifetime risk of progressing from TBI to TB disease is 5-10%, with approximately half of those who progress doing so within the first 2 years after infection [4].

Identifying the determinants of progression to TB among contacts is critical for TB control; prevention efforts could then focus on the 5-10% who would develop TB without intervention. Several studies have identified characteristics in close contacts associated with progression to TB: age (<5 years) [5], malnutrition [6], HIV infection [7], and evidence of TBI based on tuberculin skin test (TST)[8] or interferon-gamma release assay (IGRA)[9]. Conversely, TB preventive therapy (TPT) decreases the risk [8, 10]. There are also characteristics of TB source cases associated with progression to TB in their close contacts: positive sputum smear [11], increased age [12], male sex [12], and cavities on chest x-ray [13]. Factors related to decreased TB risk are household characteristics, such as access to water in pipes, or a toilet[13]. Nevertheless, these findings have varied between populations, and most of the predominant risk factors are unclear. The ability to identify contacts at high risk of progressing to TB disease would facilitate focusing TPT on these persons and thereby decrease the TB burden[2].

Previously, our group described the cascade of care among adult and child TB close contacts in Brazil [14, 15] and associations between transcriptomic signatures in whole blood and progression to active TB [16]. However, we have not evaluated risk factors for progression to TB in a prediction model. The present study developed a prediction model to identify risk factors for progressing to active TB among close contacts of pulmonary TB index cases.

## Methods

### Study Cohort and Design

This longitudinal observational study was conducted in the Regional Prospective Observational Research in Tuberculosis (RePORT)-Brazil cohort of culture-confirmed pulmonary TB cases and their close contacts. Participants were enrolled between June 2015 and June 2019 and followed for 24 months. Enrollment sites were in three Brazilian states: Rio de Janeiro (Rio de Janeiro and Caxias), Bahia (Salvador), and Amazonas (Manaus). The representativeness of the RePORT-Brazil cohort compared to all TB cases reported in Brazil has been described previously [17].

### Participants and study groups

Individuals ≥18 years old diagnosed with new or recurrent pulmonary TB (with or without extrapulmonary TB) by culture-positive sputum (Lowenstein-Jensen medium or BD BACTEC MGIT) were enrolled. After enrollment of the TB index case, close contacts identified were invited to be interviewed and examined at RePORT-Brazil healthcare units. Close contacts were defined as individuals exposed to a culture-positive pulmonary TB case for ≥4 hours in one week in the 6 months prior to TB diagnosis.

Contacts were classified into three groups according to their interferon-gamma release assay (IGRA) result at baseline and TPT status: (i) all contacts, regardless of IGRA and TPT status; (ii) contacts who were IGRA-positive at baseline, and (iii) contacts who were IGRA-positive at baseline and did not receive TPT or received TPT for <30 days. These groups differed clinically but were not mutually exclusive.

### Ethics Statement

The RePORT-Brazil protocol, informed consent, and study documents were approved by the institutional review boards at each study site and at Vanderbilt University Medical Center. Participation in RePORT-Brazil was voluntary, and written informed consent was obtained from all participants or their legally responsible guardians. All data were de-identified.

### Procedures

Enrolled contacts were investigated for TBI and evaluated in-person at baseline and 6 months after enrollment; subsequent assessments every 6 months were by telephone, then in-person if they reported signs or symptoms of TB disease. At baseline, we performed a clinical evaluation; contacts were enrolled if they reported no TB symptoms, including weight loss. Chest X-ray was performed and blood collected for IGRA and HIV testing. Clinical, laboratory, and socio-demographic data were collected through standardized case report forms in REDCap[18]. All procedures were performed according to Brazilian National TB Guidelines[19]. Contacts with a negative or indeterminate baseline IGRA test underwent repeat testing at month 6. IGRA collection, processing and interpretation were performed according to the manufacturer’s recommendations for the QuantiFERON®-TB Gold in tube assay (Qiagen) until May 2017, and the QuantiFERON®-TB Gold Plus assay subsequently.

Contacts i) with a positive IGRA result, or ii)<5 years old or iii) living with HIV (ii and iii regardless of IGRA result), were recommended to receive TPT according to Brazilian guidelines[19]. The TPT regimen was isoniazid 5-10mg/kg daily in adults and 10 mg/kg in children (300 mg daily maximum for both) for 6-9 months. Complete TPT was defined as receiving ≥6 months of isoniazid.

### Outcome

For this study, progression to TB was defined as TB among contacts confirmed a)microbiologically by a positive Xpert (MTB/RIF or Ultra) or solid (Lowenstein-Jensen) or liquid (BD MGIT) culture; or b)clinically, based on abnormal chest X-ray, suggestive histology (e.g., caseating or necrotizing granulomas) and clinical symptoms such as cough, fever, night sweats and weight loss occurring within 24 months of enrollment.

### Statistical Analysis

Characteristics of study participants were displayed as median and interquartile ranges (IQR) for continuous variables and frequency (%) for categorical variables. Continuous variables were compared using the Mann-Whitney *U* test (between two groups) or Kruskal-Wallis test (between more than two groups). Categorical variables were compared using Fisher’s exact test (2×2 comparisons) or Pearson’s chi-square test (more than two groups). Only information collected at baseline was used for analysis. P-values <0.05 were considered statistically significant.

### Dimension Reduction

Due to the low number of TB progressors among contacts and the high number of potential predictors, dimension reduction techniques were performed in the following order: (i) empirical review of potential predictor variables and (ii)least absolute shrinkage and selection operator (LASSO) regression analysis. Only variables included in the previous stage were entered into the next stage.

A set of possible predictor variables was selected a priori and identified by consensus of the co-authors. The empirical review excluded similar variables (e.g., CAGE assessment and alcohol variables) that could be adequately explained or measured by other variable(s) totally present or absent in all progressors to TB. The last stage used LASSO regression to further select the variables to be included in the final prediction model. Because of the low number of events, the magnitude of the penalty term was chosen by leave-one-out cross-validation. Each dimension reduction stage was carried out to find potential predictors for every study group in a hierarchical order.

We also conducted a secondary approach in which principal component analysis (PCA) was used as an intermediate step between the empirical review and the LASSO regression (see **Supplementary Material**). PCA was conducted to remove correlated variables by clustering predictors; components were interpreted based on moderate correlation (correlation coefficient: 0.6), either positive or negative.

### Prediction Models and validation

Three prediction models were performed using mixed-effects Cox proportional hazard models. We used a random intercept and treated TB index cases as clusters to account for possible correlation within close contacts of the same index case. Time to TB progression was calculated for each participant who progressed to TB; participants who did not progress were censored at their last visit date or the end of the study period. Variables selected for each study group after the dimension reduction step were entered into each model. Results are presented as point estimates and their corresponding 95% confidence intervals (CI).

We performed internal validation to assess the ability of the prediction models to discriminate between progressors to TB vs no progression. Discrimination was quantified with the area under the receiver operating characteristic (ROC) curve. Since the same dataset was used to construct and evaluate the performance of the three prediction models, their predictive ability may be overly optimistic. Therefore, we used 10000 bootstrapped samples to derive optimism-corrected performance measures [20].

### Participants with incomplete observations

We performed two analyses to account for missingness in the data. A complete-case analysis, in which participants with missing information on variables retained after the empirical review, were excluded from subsequent steps (e.g., excluded from the LASSO). Missingness was <1%, except for income and abnormal chest X-ray (missing for 3.4% and 10.8% of the participants, respectively). We also re-ran the analyses with an imputed dataset. Missing data were imputed using chain equations [21]. We used a single imputation, only to verify whether results would differ from the complete-case analysis.

### Critical values for clinical interest and decision-making

The third prediction model was restricted to contacts who were IGRA-positive at baseline and did not receive TPT or received TPT for <30 days. In this subgroup, due to the limited number of events, visual plots, instead of p-values, were used to assess the relationship between BMI and the odds of progressing to active TB. Although the non-linear relationship was informative for prediction accuracy, it was less informative for clinical input and decision-making. Thus, we also performed a segmented (piecewise) logistic regression model restricted to this population, to find the critical value of BMI associated with a significant change in the odds of progressing to active TB. The piecewise model was fitted assuming, for each segment, a linear relationship between BMI and the odds of having active TB; the optimal ‘breakpoint’, i.e., the point where there was a significant change in the relationship between BMI and the outcome, was estimated via maximum likelihood. The optimal breakpoint is presented as a point estimate with its corresponding 95%CI.

## Results

### Characteristics of study population

The study population included 1846 close contacts of 619 pulmonary TB cases enrolled in RePORT-Brazil (**Supplementary Figure 1**). Of 1846, 678 were IGRA-positive, 1132 were IGRA-negative, and 31 were IGRA-indeterminate at baseline. Five contacts did not have IGRA testing performed (**Supplementary Figure 2**). Among the IGRA-positive contacts, 18 progressed to TB; among IGRA-negative contacts, 6 progressed to TB. One of the contacts without IGRA testing progressed to TB (**Supplementary Figure 2**). Of the 25 contacts who developed TB, (1.4% of 1846 contacts), 13 had microbiologically confirmed TB, 2 had histopathological evidence suggestive of TB, and 10 had clinical TB. In addition, 19 (75%) had pulmonary TB and 6 (24%) had extrapulmonary TB.

No significant differences were found in the clinical characteristics of TB contacts (and their index cases) who did vs. did not progress to TB, except TB progressors were more likely to be IGRA-positive than non-progressors (75% vs. 36.3%) (**Table 1**). More details are in **Table 1**.

**Table 1.** Characteristics of TB contacts and TB cases by progression to TB status.

| Characteristics | n/N | No progression to TB (n=1821) | Progression to TB (n=25) | p-value |
| --- | --- | --- | --- | --- |
| **TB contacts** |  |  |  |  |
| **Age – median (IQR)** | 1846/1846 | 31.8 (16.1-64.9) | 31.9 (20.2-71.1) | 0.333 |
| **Female– no. (%)** | 1846/1846 | 1075 (59) | 16 (64) | 0.686 |
| **Race/Ethnicity – no. (%)** | 1844/1846 |  |  | 0.242 |
| White |  | 356 (19.6) | 6 (24) |  |
| Black |  | 363 (20) | 7 (28) |  |
| Asian |  | 5 (0.3) | 0 (0) |  |
| Pardo |  | 1081 (59.4) | 12 (48) |  |
| Indigenous |  | 14 (0.8) | 0 (0) |  |
| **BMI – median (IQR)** | 1845/1846 | 24.3 (20.2-36) | 24.2 (19.6-32.2) | 0.608 |
| **BMI- categories– no. (%)** | 1845/1846 |  |  | 0.264 |
| < 18.5 |  | 326 (17.9) | 4 (16) |  |
| 18.5 - 25 |  | 686 (37.7) | 13 (52) |  |
| > 25 |  | 807 (44.4) | 8 (32) |  |
| **Literate– no. (%)** | 1843/1846 | 1635 (89.9) | 23 (92) | 0.733 |
| **Income – no. (%)** | 1784/1846 |  |  | 0.177 |
| Without income |  | 373 (21.2) | 6 (26.1) |  |
| Equal or less than a minimum wage |  | 717 (40.7) | 12 (52.2) |  |
| More than a minimum wage |  | 671 (38.1) | 5 (21.7) |  |
| **BCG scar – no. (%)** | 1845/1846 | 1631 (89.6) | 21 (84) | 0.323 |
| **HIV infection – no. (%)** | 1844/1846 | 48 (2.6) | 0 (0) | NA |
| **ART (if HIV+) – no. (%)** | 48/48 | 38 (79.2) | 0 (0) | NA |
| **Smoking – no. (%)** | 1845/1846 |  |  | 0.204 |
| Current |  | 188 (10.3) | 1 (4) |  |
| Former |  | 296 (16.3) | 3 (12) |  |
| Never |  | 1336 (73.4) | 21 (84) |  |
| **Passive smoking – no. (%)** | 1838/1846 | 585 (32.3) | 10 (40) | 0.398 |
| **Alcohol consumption – no. (%)** | 1845/1846 |  |  | 0.121 |
| Current |  | 624 (34.3) | 6 (24) |  |
| Former |  | 350 (19.2) | 3 (12) |  |
| Never |  | 846 (46.5) | 16 (64) |  |
| **Illicit drug use – no. (%)** | 1845/1846 |  |  | 0.101 |
| Current |  | 43 (2.4) | 0 (0) |  |
| Former |  | 148 (8.1) | 0 (0) |  |
| Never |  | 1629 (89.5) | 25 (100) |  |
| **Abnormal chest X-ray– no. (%)** |  | 8 (0.4) | 0 (0) | NA |
| **Diabetes– no. (%)** | 1821/1846 | 89 (5) | 1 (4) | 1 |
| **Hypertension– no. (%)** | 1821/1846 | 219 (12.2) | 3 (12) | 1 |
| **Time index contact (weeks) – median (IQR)** | 1821/1846 | 37 (13-255) | 20 (5-144) | 0.063 |
| **TPT status – no. (%)** | 1846/1846 |  |  | 0.439 |
| Not recommended |  | 983 (54) | 4 (16) |  |
| Recommended, never started |  | 352 (19.3) | 16 (64) |  |
| Recommended, incomplete TPT |  | 168 (9.2) | 4 (16) |  |
| Recommended, complete TPT |  | 318 (17.5) | 1 (4) |  |
| **IGRA result at baseline– no. (%)** | 1841/1846 |  |  | **0.001** |
| Negative |  | 1126 (62) | 6 (25) |  |
| Positive |  | 660 (36.3) | 18 (75) |  |
| Indeterminate |  | 31 (1.7) | 0 (0) |  |
| **TB index cases** |  |  |  |  |
| **Age – median (IQR)** | 1846/1846 | 34 (24-64) | 38 (26-64) | 0.710 |
| **Female– no. (%)** | 1842/1846 | 1126 (62) | 17 (68) | 0.679 |
| **Multidrug resistance – no. (%)** | 1814/1846 | 29 (1.6) | 0 (0) | NA |
| **HbA1c (%)–median (IQR)** | 1839/1846 | 5.9 (5.5-12.1) | 5.8 (5.3-13.8) | 0.707 |
| **Cough at baseline – no. (%)** | 1842/1846 | 1682 (92.6) | 25 (100) | 0.252 |
| **Abnormal chest X-ray– no. (%)** | 1647/1846 | 833 (51.4) | 15 (60) | 0.426 |
| **Smoking – no. (%)** | 1842/1846 |  |  | 0.765 |
| Current |  | 325 (17.9) | 4 (16) |  |
| Former |  | 598 (32.9) | 8 (32) |  |
| Never |  | 894 (49.2) | 13 (52) |  |
| **Passive smoking – no. (%)** | 1830/1846 | 1190 (65.9) | 17 (68) | 1 |
| **Alcohol consumption – no. (%)** | 1842/1846 |  |  | 0.658 |
| Current |  | 680 (37.4) | 10 (40) |  |
| Former |  | 793 (43.6) | 8 (32) |  |
| Never |  | 344 (18.9) | 7 (28) |  |
| **Illicit drug use – no. (%)** | 1842/1846 |  |  | 0.287 |
| Current |  | 200 (11) | 2 (8) |  |
| Former |  | 428 (23.6) | 4 (16) |  |
| Never |  | 1189 (65.4) | 19 (76) |  |
| **Positive sputum smear – no. (%)** | 1820/1846 | 1373 (76.5) | 20 (80) | 0.815 |

**Table note.** Data represent no. (%), except for age, BMI and HbA1c, which is presented as median and interquartile range (IQR). Continuous variables were compared using the Mann Whitney U test and categorical variables were using the Pearson's chi-square test. Bold values represent statistically significant. Abbreviations: TB: tuberculosis, BMI: Body Mass Index, ART: antiretroviral therapy, IGRA; Interferon- Gamma Release Assay, TPT: Tuberculosis preventive treatment.

### Dimension Reduction

For this study, 30 variables were collected at baseline and evaluated: 19 variables of TB contacts and 11 of TB cases. After the empirical review stage, 22 variables were selected as candidate variables for the (time-to-event) LASSO regression; in this stage, five variables of contacts were retained: income, smoking, time difference between visit 1 of the TB index case and visit 1 of the contact, TPT, and positive IGRA result at baseline (**Figure 1**). The inclusion and exclusion of variables in each stage are displayed in **Supplementary Table 1.** For the PCA-LASSO, which used PCA as an intermediate step between empirical review and LASSO, three variables were retained: time index-contact, TPT, and positive IGRA test result at baseline. More details for the PCA-LASSO are displayed in **Supplementary Table 2.**

**Figure 1.**
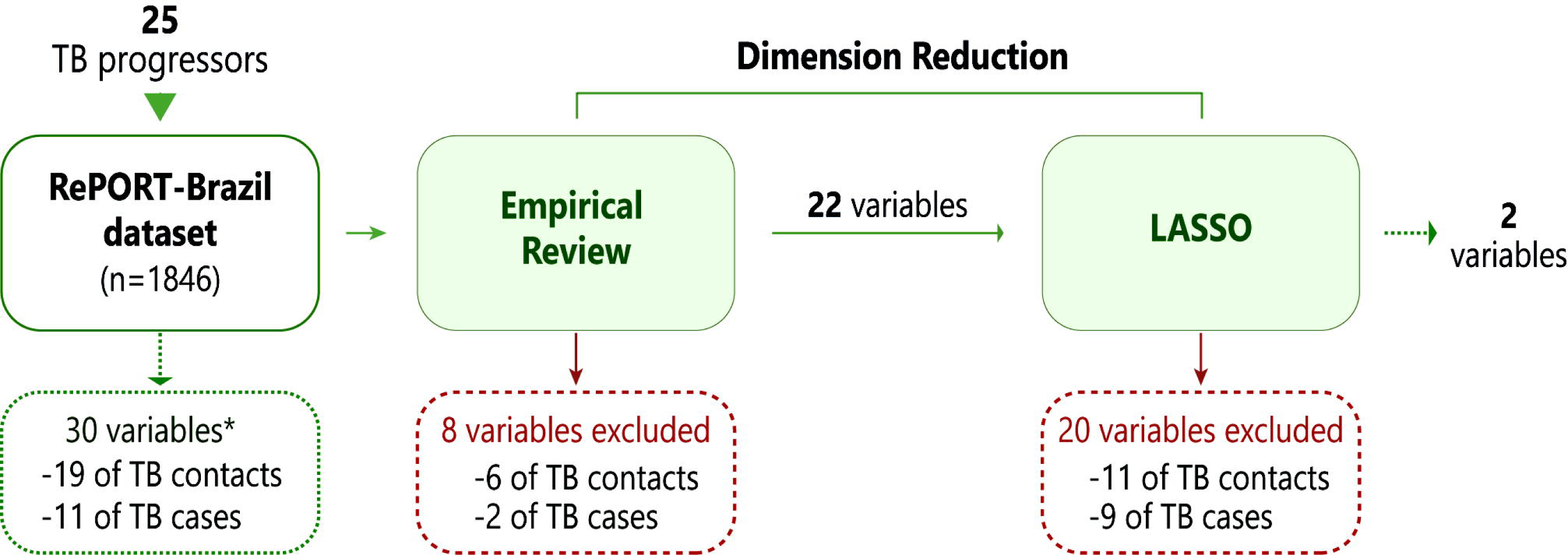
Dimension reduction stages. This scheme shows the number of variables initiated, excluded, and retained at empirical review and LASSO regression stages. More details are in **Supplementary Table 1.** *Data collected at baseline. Abbreviations: TB: Tuberculosis.

### Determinants for progression to TB among the entire TB contact population

A mixed-effects Cox proportional hazard model quantified the effect of each of the variables selected in the dimension reduction steps on the risk of progressing to active TB. The results demonstrated that of the three categories of the TPT variable (never started, incomplete, completed and not recommended), never started TPT [adjusted hazard ratio (aHR): 11.79, 95% confidence interval (CI):1.55-89.77, p-value:0.017] was associated with progression to TB (**Figure 2A**). Internal validation of the model estimated with optimism-corrected AUC of 0.85 (95%CI: 0.78–0.91) was obtained after 10000 bootstrap replications (**Figure 2B**).

**Figure 2.**
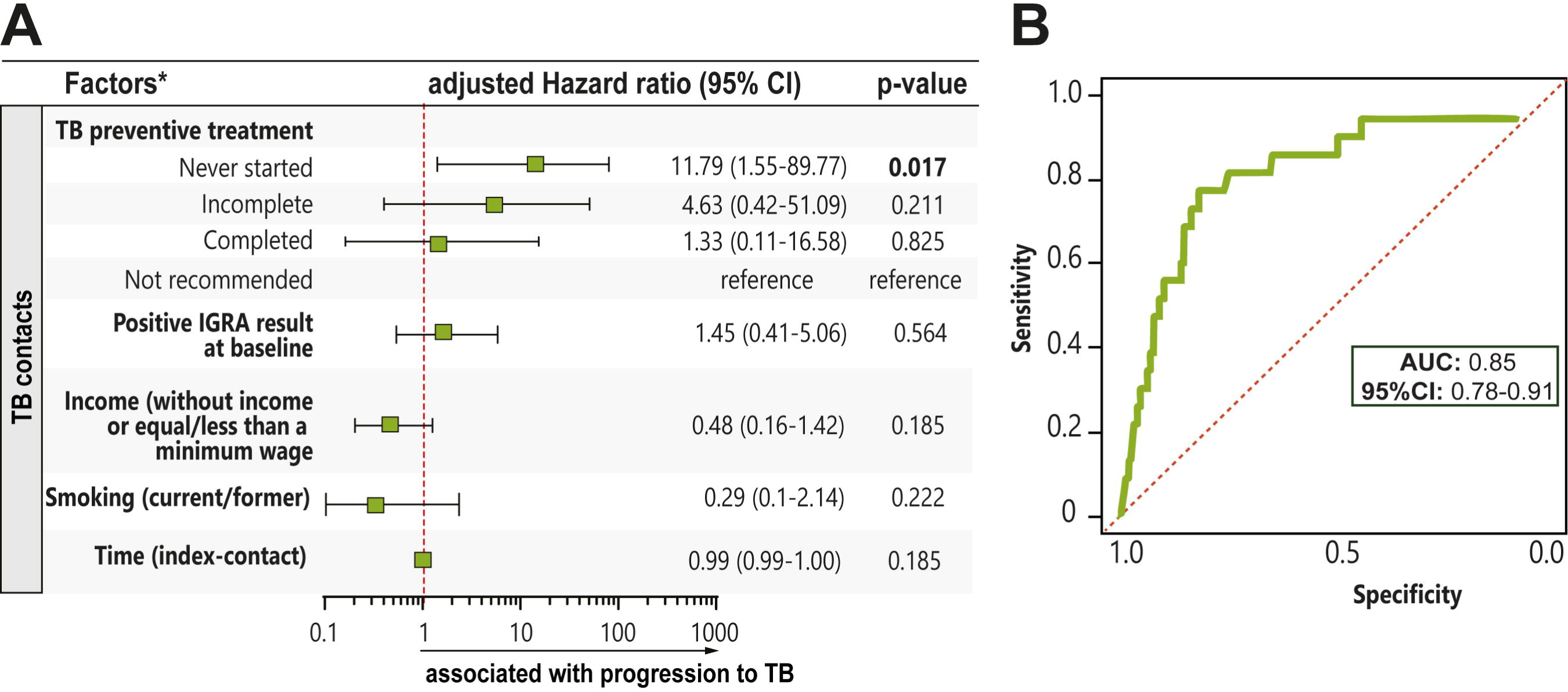
**(A)** Mixed-effects Cox proportional hazard model was performed to test association between characteristics of TB contacts or pulmonary TB cases and progression to TB among all close contacts enrolled in the study. Dimension reduction was used to select characteristics of TB contacts and TB cases included in this model. **(B)** Receiver operator characteristics (ROC) curve for model development using 10000 bootstraps. More details are in **Table 1** and **Supplementary Table 1.** *Data collected at baseline. Abbreviations: IGRA: Interferon-Gamma Release Assay, TB: Tuberculosis, TPT: Tuberculosis preventive treatment, Time (index-contact): time difference between visit 1 of the TB index case and visit 1 of the contact, AUC: area under the curve, CI: confidence interval.

### TB progression among contacts who were IGRA-positive at baseline

We then evaluated progression to TB among persons who were IGRA-positive at baseline. Of 678 contacts, 18 progressed to TB. The univariable analysis showed that 77.8% of TB progressors never started TPT and only 5.6% completed TPT (p<0.001). No other significant differences were found in the characteristics of these groups. (**Supplementary Table 3**). Dimension reduction employing empirical review-LASSO (**Supplementary Table 4**) were performed; of 29 initial variables, only TPT was retained for the final model. Analysis using a mixed-effects Cox model with the selected variables showed that never started TPT was associated with progression to TB (aHR:13.12, 95%CI:1.72-99.75, p-value:0.013) (**Figure 3**). The optimism-corrected AUC of this model was 0.74 (95%CI: 0.65–0.80). Using PCA-LASSO for variable selection (**Supplementary Table 5)**, TPT and characteristics of TB index cases as age, secondary smoking and drug use were selected.

**Figure 3.**
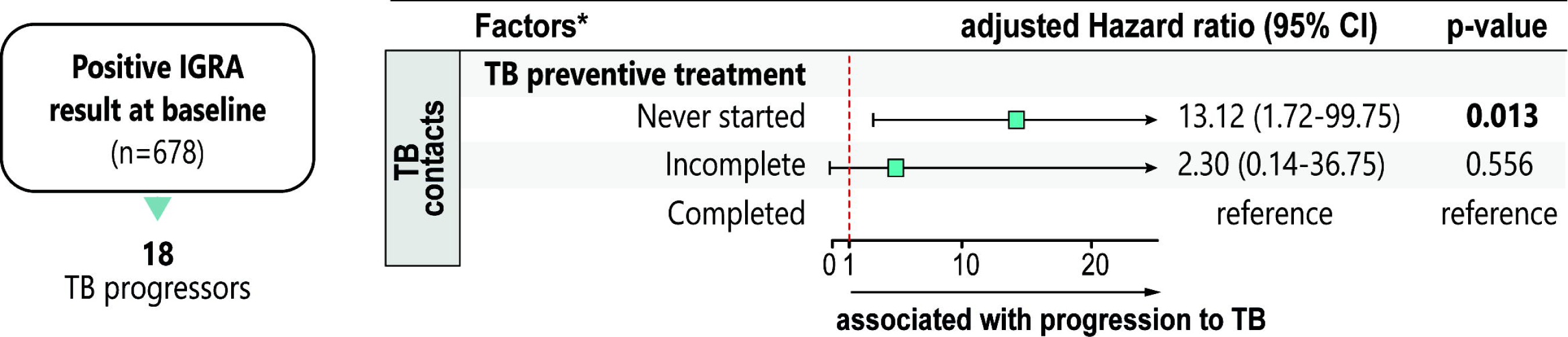
Mixed-effects Cox proportional hazard model was performed to test the association between characteristics of TB contacts or pulmonary TB cases and progression to TB among contacts with positive IGRA result at baseline. Dimension reduction was used to select characteristics of TB contacts and TB cases included in this model. More details are in **Supplementary Table 3 and 4.** *Data collected at baseline. Abbreviations: IGRA: Interferon-Gamma Release Assay, TB: Tuberculosis, TPT: Tuberculosis preventive treatment, CI: confidence interval.

### Progression to TB among contacts who were IGRA-positive and did not receive TPT or received TPT for <30 days

We then assessed the study population of greatest interest: the 285 contacts who were IGRA-positive at baseline but did not start TPT or received TPT for <30 days. Of the 285, 15 progressed to TB disease. When comparing characteristics of these contacts by progression to TB status, we found that the median BMI of TB progressors [24.2 kg/m^2^ (IQR:20.4-28.1)] was significantly lower than that of non-progressors [25.3 kg/m^2^ (IQR: 22.1-36.6)] (p-value:0.04) (**Supplementary Table 6**). Dimension reduction employing empirical review-Lasso reduced from 28 to one potential predictors: BMI of contacts (**Supplementary Table 7**) and PCA-Lasso: BMI and passive smoking in TB index cases (**Supplementary Table 8**). Using these variables in the mixed-effects Cox regression model, we found that for each unit increase of BMI, the risk of progressing to TB decreased by 13% (aHR:0.87, 95%CI:0.78-0.98, p-value:0.020) (**Figure 4**). The optimism-corrected AUC of this model was 0.67 (95%CI 0.56–0.78).

**Figure 4.**
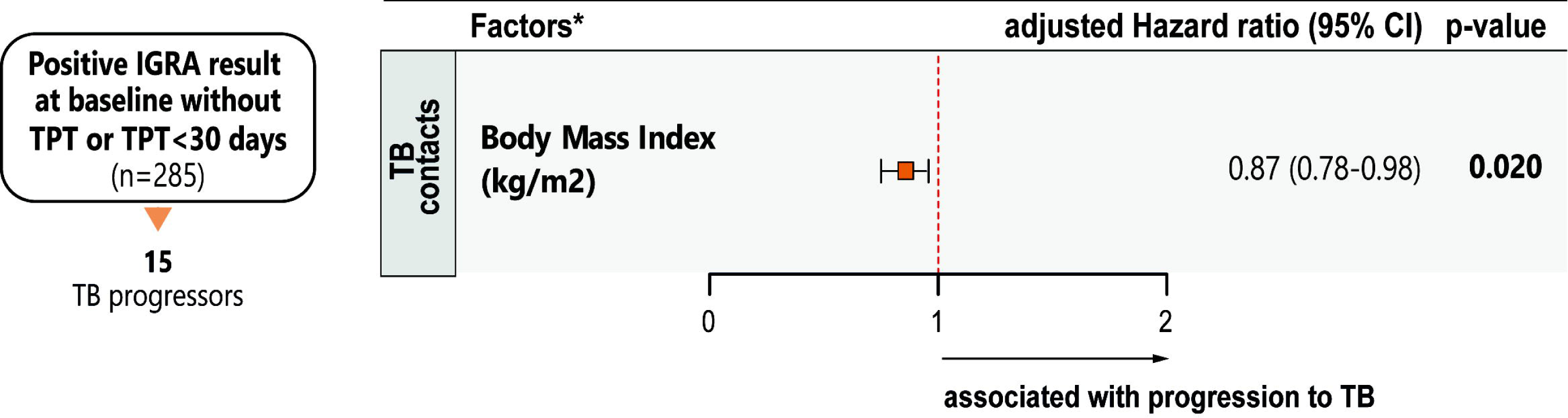
Mixed-effects Cox proportional hazard model was performed to test associations between characteristics of TB contacts or pulmonary TB index cases and progression to TB among contacts with a positive IGRA result at baseline who did not start treatment or received <30 days. Dimension reduction was used to select the variables included in this model. More details are in **Supplementary Table 6 and 7.** *Data collected at baseline. Abbreviations: IGRA: Interferon-Gamma Release Assay, TB: Tuberculosis, CI: confidence interval.

We performed additional analyses to identify the critical value of BMI that led to the highest change in the odds of progressing to active TB. Through a segmented (piecewise) logistic regression analysis, the optimal breakpoint of 24.93 kg/m2 [95%CI: (20.25-29.61)] was estimated **(Figure 5A).** Using this critical point, we found that those contacts with BMI <25 kg/m^2^ were more likely to progress to active TB [aHR:4.14 (95%CI:1.17-14.67, p-value:0.028)] **(Figure 5B).** Internal validation of this model estimated an AUC of 0.66 (95%CI: 0.54–0.72).

**Figure 5.**
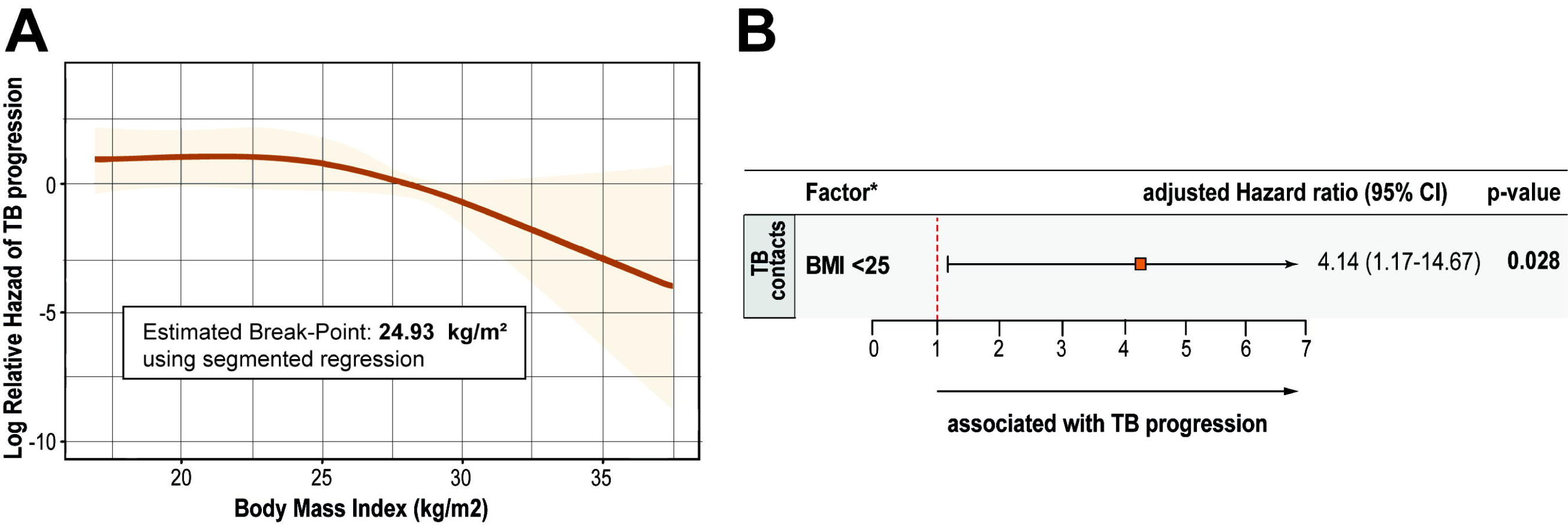
**(A)** Regression Model with Segmented Relationship to find the optimal break-point using BMI values of contacts with IGRA-positive result at baseline and who did not start treatment or received <30 days. **(B)** Logistic regression using the optimal break-point BMI among contacts with positive IGRA result at baseline and who did not start treatment or received <30 days. BMI ≥ 25 kg/m^2^ was used as the reference. *Data collected at baseline. Abbreviations: IGRA: Interferon-Gamma Release Assay, TB: Tuberculosis, CI: confidence interval.

Moreover, we calculated the probability of progressing to active TB in the group of contacts who were IGRA-positive at baseline and did not receive TPT or received TPT for <30 days: 5.26% (15 progressors/285 contacts). Among IGRA-positive contacts with BMI <25 kg/m^2^ who did not receive TPT, the TB risk was 8.4% (12 progressors/143 contacts); in such contacts with BMI ≥25 kg/m^2^ the probability was 2.1% (3 progressors/141 contacts). In addition, the breakpoint of 18.5 kg/m² was not associated with progression to TB (HR:1.23, 95%CI:0.39-3.92), using the breakpoint 25 kg/m² as the reference level.

Likewise, when we used the variables selected through dimension reduction by empirical review, PCA and LASSO regression in the models for each study group, the predictors significantly associated with progression to TB were the same as those found when we used dimension reduction by empirical review-LASSO regression (**Supplementary Table 9, 10 and 11)**. Also, results for the imputed analyses were similar to the complete-case in each group of the study and are presented in **Supplementary Table 12, 13 and 14**.

## Discussion

We found that close TB contacts who were IGRA-positive at baseline but did not receive TPT and had lower BMI (i.e.,<25 kg/m^2^) were at increased risk of progression to TB. Of note, 12 of 15 (80%) of those who progressed to TB were identified by this algorithm. Previous studies have reported underweight as a risk factor for incident active TB [22, 23] while individuals with overweight and normal weight had a reduced risk of TB disease [24]. In a recent post-hoc analysis of a clinical trial among people with HIV treated with TPT (not close contacts; 38% were IGRA-positive), BMI <25 kg/m^2^ was also associated with increased TB risk [25, 26]. Our study population of IGRA-positive close TB contacts who did not receive TPT (none of whom had HIV) is particularly relevant. This BMI value coincides with the upper value for the normal BMI classification (18-25 kg/m^2^).

Previous studies have shown that overweight and obese individuals (though not specifically close contacts of people with TB) have a lower risk of TB, compared to those with normal BMI and low BMI [27, 28]. Although the impact of immunological, biological and genetic mechanisms of BMI on TB progression is complex, nutritional differences between overweight or obese individuals and those with normal or low weight may be one of the possible factors explaining the protective effect against TB progression [27].

Low BMI can influence innate and adaptive immune responses through various mechanisms [29]. There is evidence of a correlation between low BMI in people with pulmonary TB and decreased levels of IFNγ, TNFα, IL-2, IL-17, IL-6, and IL-12, affecting protective immunity [29]. Reduced chemokine levels have also been observed in individuals with TB infection [29]. Moreover, Mtb can use adipose tissue to survive without being recognized by the host immune system [30], which would explain why overweight or obesity are protective factors for the development of TB [30]. Corroborating this, a study of cost-effectiveness models for reducing TB incidence and mortality in India showed that a robust nutritional intervention could prevent 78% of TB cases and 88% of deaths among malnourished close contacts [31]. Furthermore, a clinical trial also in India demonstrated a reduced incidence of pulmonary TB (48%) among household contacts of TB cases who received nutritional support [32].

Using characteristics of close TB contacts and their index cases, we performed a prediction model for progression to TB at the time of evaluation of the close contact. Among close contacts with Mtb infection (i.e., IGRA-positive), BMI was the only variable that predicted progression to TB. Among IGRA-positive close contacts who did not receive preventive therapy, BMI<25 was associated with an approximately 4-fold increased odds of progressing to TB. In addition, the number of contacts needing treatment decreased from 285 to 143, a 50% reduction.

In the prediction model among all close contacts, as well as among IGRA-positive close contacts, no TPT start was the strongest risk factor for progression to TB. TPT is highly effective in preventing progression to TB disease [33, 34]. Two studies, one in Pakistan [34] and another using a decision analysis model [33] reported that contact investigation programs related to the initiation of TPT prevent progression to TB disease compared to no intervention, varying the number of cases averted according to the type of contact (age and type of exposure), TPT scheme and detection algorithm [33, 34].

In 2018, WHO made a conditional recommendation to provide TPT to all household contacts of all ages [35], with many countries expanding the provision of TPT from children <5 years to children <15 years regardless of IGRA result. Despite these recommendations, in 2021, only 40% of the TPT household contacts aged <5 years and 3% of contacts ≥5 years received TPT [36]. Thus, TPT implementation remains deficient among contacts of all ages.

In the full cohort (regardless of IGRA and TPT status) TB incidence among contacts was 1.4%. A recent systematic review [37] found an incidence of active TB among household contacts of 2% in the first year and 0.75% in the second year following exposure; incidence declined in subsequent years, regardless of IGRA and TPT status. Among close contacts in this study who were IGRA-positive and did not receive TPT, 5% progressed to TB. This is consistent with several previous studies [38, 39].

This study had several limitations. First, we did not obtain information on the type of contacts (e.g., household, coworkers) and relevant comorbidities (e.g., diabetes). This could have influenced an over-or under-fitting in the statistical model. Second, it was not possible to determine whether the contact became infected after exposure or if there was pre-existing Mtb infection. Third, the number of TB progressors was low; however, the proportion of progressors was consistent with the existing literature: 1.4% of all contacts and 5% of IGRA-positive contacts who did not receive preventive therapy [38, 39]. Fourth, the proportion of study participants with BMI<18.5 kg/m2 was relatively low (330/1845; 17.9%), and cultures were not obtained in individuals without symptoms. However, none of the TB progressors reported weight loss or had an abnormal chest X-ray at baseline, making it less likely that they had undiagnosed TB. Finally, external validation of the main prediction model could not be performed. Confirmation of this BMI-based approach, and the BMI cutoff value of 25 kg/m^2^, in a separate cohort would provide important evidence before putting this into clinical practice. However, the dimension reduction approach at each step, from the selection of variables to the final mixed-effects Cox proportional hazard analysis, was exhaustive, and we achieved robust internal validation in a large, diverse cohort.

In conclusion, BMI<25 kg/m^2^ identified IGRA-positive close TB contacts who were at high risk of progressing to TB disease. TPT should be targeted to this high-risk group to maximize TB prevention efforts.

## NOTES

## Acknowledgements

The authors thank the study participants. We also thank the clinical and laboratory teams of RePORT Brazil.

## Disclaimer

The funders had no role in the study design, data collection and analysis, decision to publish, or preparation of the manuscript.

## Data availability

The data used in the manuscript is available upon request to the corresponding author.

## Contributions

Conceptualization, M.C.F., M.C.S., V.C.R., B.B.A and T.R.S..; Data curation, M.B.A., G.A., M.C.F., C.S. and T.R.S.; Investigation, M.B.A., G.A, M.C.F., C.S., and T.R.S.; Formal analysis, M.B.A., P.F.R. and G.A.; Funding acquisition, A.L.K., M.C.S., M.C.F., V.C.R., B.B.A and T.R.S.; Methodology, M.B.A., G.A., and T.R.S.; Project administration, M.C.F., C.S and T.R.S.; Resources, M.B.A., G.A., M.C.F., C.S. and T.R.S.; Software, M.B.A., G.A., and T.R.S.; Supervision, P.F.R. and T.R.S.; Writing—original draft, M.B.A., G.A., P.F.R. and T.R.S.; Writing review and editing, all authors. All authors have read and agreed to the submitted version of the manuscript.

