## Supplementary Appendix for "Body Mass Index and Incident Tuberculosis in Close Tuberculosis Contacts"

[Supplementary Table 6 . Characteristics of TB contacts with positive IGRA result at baseline and did not start treatment or received <30 days and TB index cases by TB progression status.	12](#_Toc190959065)

[Supplementary Table 7. Characteristics of TB contacts with positive IGRA result at baseline and did not start treatment or received <30 days and TB cases included and excluded in each Dimension Reduction stage using empirical review and LASSO regression.	14](#_Toc190959066)

[Supplementary Table 8. Characteristics of TB contacts with positive IGRA result at baseline and did not start treatment or received <30 days and TB cases included and excluded in each Dimension Reduction stage using empirical review, PCA and LASSO regression.	15](#_Toc190959067)

[Supplementary Table 11. Mixed-effects Cox proportional hazard model for progression to TB among close contacts with a positive IGRA result at baseline who did not start treatment or received <30 days enrolled in the study.	16](#_Toc190959070)

[Supplementary Table 14. Mixed-effects Cox proportional hazard model for progression to TB among close contacts with a positive IGRA result at baseline who did not start treatment or received <30 days enrolled in the study with imputed data.	18](#_Toc190959073)

### Supplementary methods

### Dimension Reduction with Principal Components Analysis

The rationale for using principal components analysis (PCA) as a secondary dimension reduction technique was to allow us to more carefully target variables of potential clinical or mechanistic interest as candidates for the LASSO. For example, variables that could help explain the variability of the data. The main goal was to develop a prediction model, not perform statistical inference. Inference after variable selection is not straightforward, as the 95% CIs for parameter estimates are generally not reliable. This is because in this two-step procedure we look at the data twice: once to select the variables, and then again to fit the regression model and run hypotheses tests, leading to CIs that may be too narrow.(1) However, some recent work suggests that this two-step approach could lead to correct standard errors and correct p-values for variables selected using the LASSO, leading to valid inference (2) . This result is asymptotical, though, and inference from studies with small sample sizes (e.g., few number of events) such as ours need to be interpreted with caution. Nevertheless, we decided to retain CIs and p-values for completeness, stressing once again that the main goal was *prediction*, while acknowledging its limitation for making inference.

### Supplementary Figures

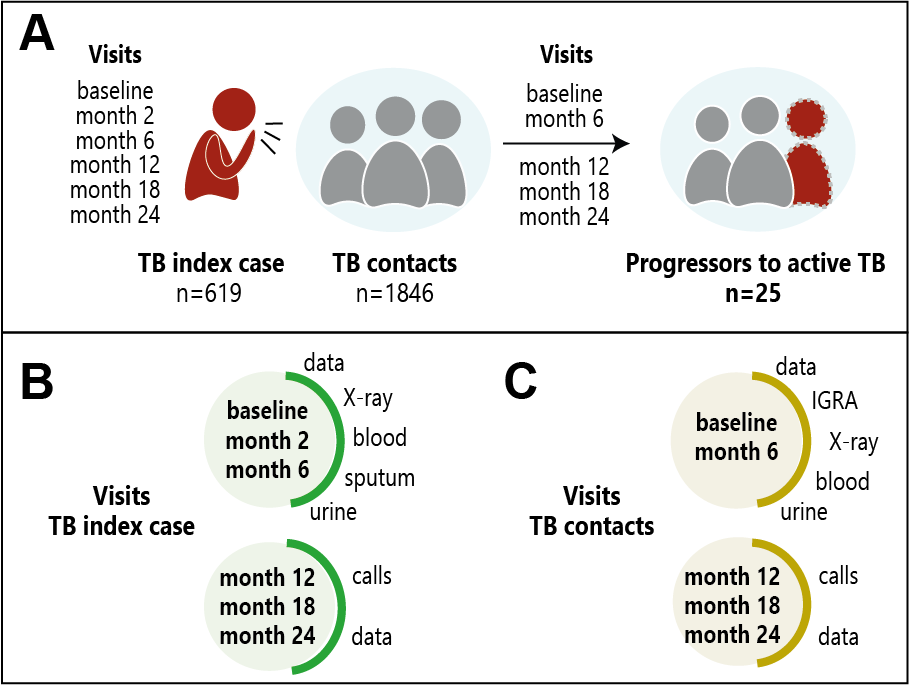

Supplementary Figure 1. Study procedures. **(A)** The number of TB index cases and TB contacts enrolled. **(B)** Scheme of the procedures at each visit for TB index cases and **(C)** TB contacts.

There were 4,145 close contacts referred from 1,187 pulmonary TB cases; of these, 2,483 contacts of 641 culture confirmed pulmonary TB cases were reached, and invited to health care units for Mtb infection screening. Of the 2,483 contacts who were invited to participate in the study, 1846 (75%) agreed to participate, provided informed consent, and were enrolled in the study.

Abbreviations: IGRA: Interferon-Gamma Release Assay, TB: Tuberculosis, TPT: Tuberculosis preventive treatment.

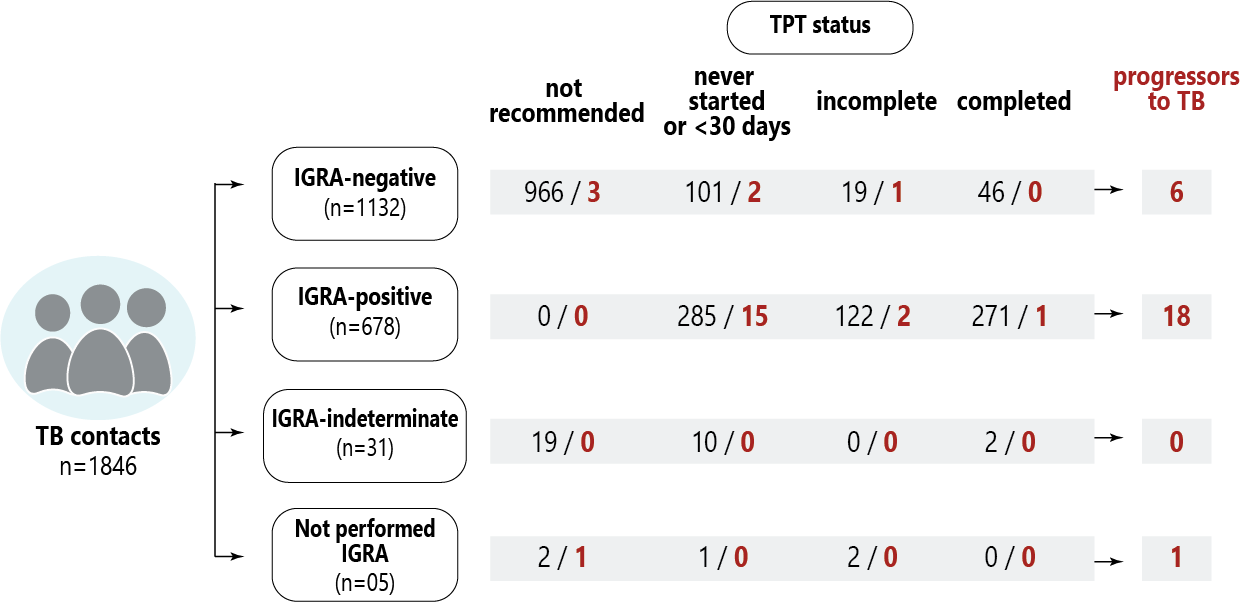

Supplementary Figure 2. Study flowchart. This figure shows the number of contacts enrolled in the study and the number of who progressed to active TB by IGRA results at baseline and TPT status.

Abbreviations: IGRA: Interferon-Gamma Release Assay, TB: Tuberculosis, TPT: Tuberculosis preventive treatment.

### Supplementary Tables

Supplementary Table 1. Characteristics of study population included and excluded in each Dimension Reduction stage using empirical review and LASSO regression.

| **Characteristics** | | **Dimensional Reduction** | |
| --- | --- | --- | --- |
|  |  | **Empirical Review*** | **LASSO** |
| **TB contact** | Age | Included | **Excluded** |
|  | Sex | Included | **Excluded** |
|  | Race/Ethnicity | Included | **Excluded** |
|  | Body Mass Index | Included | **Excluded** |
|  | Literate | Included | **Excluded** |
|  | Income | Included | Included |
|  | BCG scar | Included | **Excluded** |
|  | HIV infection | **Excluded** |  |
|  | Antiretroviral therapy (if HIV+) | **Excluded** |  |
|  | Smoking | Included | Included |
|  | Passive smoking | Included | **Excluded** |
|  | Alcohol consumption | Included | **Excluded** |
|  | CAGE assessment | **Excluded** |  |
|  | drug use | **Excluded** |  |
|  | Diabetes | **Excluded** |  |
|  | Hypertension | **Excluded** |  |
|  | Time (index-contact) | Included | Included |
|  | Tuberculosis preventive treatment | Included | Included |
|  | IGRA result at baseline | Included | Included |
| **TB cases** | Age | Included | **Excluded** |
|  | Sex | Included | **Excluded** |
|  | Multidrug resistance | **Excluded** |  |
|  | HbA1c | Included | **Excluded** |
|  | Enrollment Cough | **Excluded** |  |
|  | Cavity on chest X-ray | Included | **Excluded** |
|  | Passive smoking | Included | **Excluded** |
|  | Smoking | Included | **Excluded** |
|  | Alcohol consumption | Included | **Excluded** |
|  | Drug use | Included | **Excluded** |
|  | Positive sputum smear | Included | **Excluded** |

**Table note.** Only variables included in the previous stage were entered into the next stage.

*Eight variables were excluded: HIV infection and antiretroviral therapy (no progressors to TB were HIV+), CAGE assessment ("alcohol consumption" variable was similar), illicit drug use (no progressors to TB used illicit drugs), hypertension and diabetes (were self-reported variables), multidrug resistance (there was no TB index of progressors to TB with multidrug resistance) and cough at baseline (all TB index had cough at baseline).

Abbreviations: IGRA: Interferon-Gamma Release Assay, TB: Tuberculosis.

### **Supplementary Table 2.** Characteristics of study population included and excluded in each Dimension Reduction stage using empirical review, PCA and LASSO regression.

| **Characteristics** | | **Dimensional Reduction** | | |
| --- | --- | --- | --- | --- |
|  |  | **Empirical Review*** | **PCA** | **LASSO** |
| **TB contact** | Age | Included | **Excluded** |  |
|  | Sex | Included | Included | **Excluded** |
|  | Race/Ethnicity | Included | **Excluded** |  |
|  | Body Mass Index | Included | Included | **Excluded** |
|  | Literate | Included | **Excluded** |  |
|  | Income | Included | **Excluded** |  |
|  | BCG scar | Included | Included | **Excluded** |
|  | HIV infection | **Excluded** |  |  |
|  | Antiretroviral therapy (if HIV+) | **Excluded** |  |  |
|  | Smoking | Included | **Excluded** |  |
|  | Passive smoking | Included | **Excluded** |  |
|  | Alcohol consumption | Included | **Excluded** |  |
|  | CAGE assessment | **Excluded** |  |  |
|  | drug use | **Excluded** |  |  |
|  | Diabetes | **Excluded** |  |  |
|  | Hypertension | **Excluded** |  |  |
|  | Time (index-contact) | Included | Included | Included |
|  | Tuberculosis preventive treatment | Included | Included | Included |
|  | IGRA result at baseline | Included | Included | Included |
| **TB cases** | Age | Included | **Excluded** |  |
|  | Sex | Included | **Excluded** |  |
|  | Multidrug resistance | **Excluded** |  |  |
|  | HbA1c | Included | **Excluded** |  |
|  | Enrollment Cough | **Excluded** |  |  |
|  | Cavity on chest X-ray | Included | Included | **Excluded** |
|  | Passive smoking | Included | Included | **Excluded** |
|  | Smoking | Included | **Excluded** |  |
|  | Alcohol consumption | Included | **Excluded** | **Excluded** |
|  | Drug use | Included | Included | **Excluded** |
|  | Positive sputum smear | Included | Included | **Excluded** |

**Table note.** Only variables included in the previous stage were entered into the next stage.

*Eight variables were excluded: HIV infection and antiretroviral therapy (no progressors to TB were HIV+), CAGE assessment ("alcohol consumption" variable was similar), illicit drug use (no progressors to TB used illicit drugs), hypertension and diabetes (were self-reported variables), multidrug resistance (there was no TB index of progressors to TB with multidrug resistance) and cough at baseline (all TB index had cough at baseline).

Abbreviations: Time (index -contact): time difference between visit 1 of the TB index case and visit 1 of the contact, PCA: Principal Component Analysis, IGRA: Interferon-Gamma Release Assay, TB: Tuberculosis.

### **Supplementary Table 3.** Characteristics of TB contacts with positive IGRA results at baseline and TB index cases, by TB progression status.

| Characteristics | n/N | No progression to TB (n=660) | Progression to TB (n=18) | p-value |
| --- | --- | --- | --- | --- |
| TB contacts |  |  |  |  |
| Age – median (IQR) | 678/678 | 36.1 (18.9-68.1) | 35.6 (20.2-71.1) | 0.930 |
| Female– no. (%) | 678/678 | 419 (63.5) | 12 (66.7) | 0.497 |
| Race/Ethnicity – no. (%) | 677/678 |  |  | 0.429 |
| White |  | 109 (16.5) | 4 (22.2) |  |
| Black |  | 163 (24.7) | 5 (27.8) |  |
| Asian |  | 4 (0.6) | 0 (0) |  |
| Pardo |  | 378 (57.4) | 9 (50) |  |
| Indigenous |  | 5 (0.8) | 0 (0) |  |
| BMI – median (IQR) | 677/678 | 24.9 (21.1-36.7) | 24.2 (20.4-36.2) | 0.244 |
| BMI- categories– no. (%) | 677/678 |  |  | 0.349S3 |
| < 18.5 |  | 94 (14.3) | 3 (16.7) |  |
| 18.5 - 25 |  | 315 (47.9) | 5 (27.8) |  |
| > 25 |  | 249 (37.8.4) | 10 (55.6) |  |
| Literate– no. (%) | 677/678 | 605 (91.8) | 17 (94.4) | 1 |
| Income – no. (%) | 655/678 |  |  | 0.156 |
| Without income |  | 131 (20.5) | 5 (31.3) |  |
| Equal or less than a minimum wage | | 290 (45.4) | 8 (50) |  |
| More than a minimum wage |  | 218 (34.1) | 3 (18.8) |  |
| BCG scar – no. (%) | 677/678 | 583 (88.5) | 16 (88.9) | 1 |
| HIV infection – no. (%) | 678/678 | 11 (1.7) | 0 (0) | NA |
| ART (if HIV+) – no. (%) | 11/11 | 7 (63.6) | 0 (0) | NA |
| Smoking – no. (%) | 677/678 |  |  | 0.108 |
| Current |  | 91 (13.8) | 1 (5.6) |  |
| Former |  | 110 (16.7) | 1 (5.6) |  |
| Never |  | 458 (69.5) | 16 (88.9) |  |
| Passive smoking – no. (%) | 674/678 | 234 (35.7) | 7 (38.9) | 0.806 |
| Alcohol consumption – no. (%) | 677/678 |  |  | 0.069 |
| Current |  | 229 (34.7) | 4 (22.2) |  |
| Former |  | 127 (19.3) | 1 (5.6) |  |
| Never |  | 303 (46) | 13 (72.2) |  |
| drug use – no. (%) | 677/678 |  |  | NA |
| Current |  | 16 (2.4) | 0 (0) |  |
| Paste |  | 54 (8.2) | 0 (0) |  |
| Never |  | 589 (89.4) | 18 (100) |  |
| Diabetes– no. (%) | 671/678 | 31 (4.7) | 0 (0) | NA |
| Hypertension– no. (%) | 671/678 | 95 (14.5) | 2 (11.1) | 1 |
| Time _index-contact_ (weeks) – median (IQR) | 671/678 | 35 (9-255) | 24 (5-157) | 0.361 |
| TPT status – no. (%) | 678/678 |  |  | **<0.001** |
| Recommended; never started |  | 247 (37.4) | 14 (77.8) |  |
| Recommended; incomplete TPT |  | 143 (21.7) | 3 (16.7) |  |
| Recommended; completed TPT |  | 270 (40.9) | 1 (5.6) |  |
| TB index cases |  |  |  |  |
| Age – median (IQR) | 678/678 | 33 (24-66) | 39 (22-68) | 0.502 |
| Female– no. (%) | 675/678 | 404 (61.5) | 12 (66.7) | 0.808 |
| Multidrug resistance – no. (%) | 670/678 | 8 (1.2) | 0 (0) | NA |
| HbA1c (%)-median (IQR) | 673/678 | 5.9 (5.4-12.2) | 5.9 (5.5-13.8) | 0.948 |
| Enrollment Cough – no. (%) | 675/678 | 621 (94.5) | 18 (100) | 0.617 |
| Chest X-ray– no. (%) | 621/678 | 373 (61.9) | 12 (66.7) | 0.808 |
| Smoking – no. (%) | 675/678 |  |  | 0.671 |
| Current |  | 146 (22.2) | 3 (16.7) |  |
| Former |  | 209 (31.8) | 6 (33.3) |  |
| Never |  | 302 (46) | 9 (50) |  |
| Passive smoking – no. (%) | 673/678 | 404 (61.7) | 13 (72.2) | 0.464 |
| Alcohol consumption – no. (%) | 675/678 |  |  | 0.570 |
| Current |  | 269 (40.9) | 7 (38.9) |  |
| Former |  | 259 (39.4) | 6 (33.3) |  |
| Never |  | 129 (19.6) | 5 (27.8) |  |
| drug use – no. (%) | 675/678 |  |  | 0.125 |
| Current |  | 96 (14.6) | 1 (5.6) |  |
| Former |  | 142 (21.6) | 2 (11.1) |  |
| Never |  | 419 (63.8) | 15 (83.3) |  |
| Positive sputum smear – no. (%) | 663/678 | 529 (82) | 15 (83.3) | 1 |

**Table note.** Data represent no. (%), except for age and BMI, which is presented as median and interquartile range (IQR). Continuous variables were compared using the Mann Whitney U test and categorical variables were using the Pearson’s chi-square test. Bold values represent statistically significant.

Abbreviations: TB: tuberculosis, BMI: Body Mass Index, ART: antiretroviral therapy, IGRA; Interferon- Gamma Release Assay, TPT: Tuberculosis preventive treatment.

### **Supplementary Table 4** . Characteristics of TB contacts with positive IGRA results at baseline and TB index cases included and excluded in each Dimension Reduction stage using empirical review and LASSO regression.

| **Characteristics** | | **Dimensional Reduction** | |
| --- | --- | --- | --- |
|  |  | **Empirical Review** | **LASSO** |
| **TB contact** | Age | Included | **Excluded** |
|  | Sex | Included | **Excluded** |
|  | Race/Ethnicity | Included | **Excluded** |
|  | Body Mass Index | Included | **Excluded** |
|  | Literate | Included | **Excluded** |
|  | Income | Included | **Excluded** |
|  | BCG scar | Included | **Excluded** |
|  | HIV infection | **Excluded** |  |
|  | Antiretroviral therapy (if HIV+) | **Excluded** |  |
|  | Smoking | Included | **Excluded** |
|  | Passive smoking | Included | **Excluded** |
|  | Alcohol consumption | Included | **Excluded** |
|  | CAGE assessment | **Excluded** |  |
|  | drug use | **Excluded** |  |
|  | Diabetes | **Excluded** |  |
|  | Hypertension | **Excluded** |  |
|  | Time (index-contact) | Included | **Excluded** |
|  | Tuberculosis preventive treatment | Included | Included |
| **TB index cases** | Age | Included | **Excluded** |
|  | Sex | Included | **Excluded** |
|  | Multidrug resistance | **Excluded** |  |
|  | HbA1c | Included | **Excluded** |
|  | Enrollment Cough | **Excluded** |  |
|  | Cavity on chest X-ray | Included | **Excluded** |
|  | Passive smoking | Included | **Excluded** |
|  | Smoking | Included | **Excluded** |
|  | Alcohol consumption | Included | **Excluded** |
|  | Drug use | Included | **Excluded** |
|  | Positive sputum smear | Included | **Excluded** |

**Table note.** Only variables included in the previous stage were entered into the next stage.

*Eight variables were excluded: HIV infection and antiretroviral therapy (no progressors to TB were HIV+), CAGE assessment ("alcohol consumption" variable was similar), drug use (no progressors to TB used illicit drugs), hypertension and diabetes (were self-reported variables), multidrug resistance (there was no TB index of progressors to TB with multidrug resistance) and cough at baseline (all TB index had cough at baseline).

Abbreviatures: Time (index -contact): time difference between visit 1 of the TB index case and visit 1 of the contact, IGRA: Interferon-Gamma Release Assay, TB: Tuberculosis.

### **Supplementary Table 5.** Characteristics of TB contacts with positive IGRA results at baseline and TB index cases included and excluded in each Dimension Reduction stage using empirical review, PCA and LASSO regression.

| **Characteristics** | | **Dimensional Reduction** | | |
| --- | --- | --- | --- | --- |
|  |  | **Empirical Review** | **PCA** | **LASSO** |
| **TB contact** | Age | Included | Included | **Excluded** |
|  | Sex | Included | Included | **Excluded** |
|  | Race/Ethnicity | Included | **Excluded** |  |
|  | Body Mass Index | Included | **Excluded** |  |
|  | Literate | Included | **Excluded** |  |
|  | Income | Included | Included | **Excluded** |
|  | BCG scar | Included | Included | **Excluded** |
|  | HIV infection | **Excluded** |  |  |
|  | Antiretroviral therapy (if HIV+) | **Excluded** |  |  |
|  | Smoking | Included | **Excluded** |  |
|  | Passive smoking | Included | Included | **Excluded** |
|  | Alcohol consumption | Included | **Excluded** |  |
|  | CAGE assessment | **Excluded** |  |  |
|  | drug use | **Excluded** |  |  |
|  | Diabetes | **Excluded** |  |  |
|  | Hypertension | **Excluded** |  |  |
|  | Time (index-contact) | Included | Included | **Excluded** |
|  | Tuberculosis preventive treatment | Included | Included | Included |
| **TB index cases** | Age | Included | Included | **Excluded** |
|  | Sex | Included | **Excluded** |  |
|  | Multidrug resistance | **Excluded** |  |  |
|  | HbA1c | Included | Included | **Excluded** |
|  | Enrollment Cough | **Excluded** |  |  |
|  | Cavity on chest X-ray | Included | Included | **Excluded** |
|  | Passive smoking | Included | Included | **Excluded** |
|  | Smoking | Included | Included | **Excluded** |
|  | Alcohol consumption | Included | **Excluded** |  |
|  | Drug use | Included | Included | **Excluded** |
|  | Positive sputum smear | Included | Included | **Excluded** |

**Table note.** Only variables included in the previous stage were entered into the next stage.

*Eight variables were excluded: HIV infection and antiretroviral therapy (no progressors to TB were HIV+), CAGE assessment ("alcohol consumption" variable was similar), drug use (no progressors to TB used illicit drugs), hypertension and diabetes (were self-reported variables), multidrug resistance (there was no TB index of progressors to TB with multidrug resistance) and cough at baseline (all TB index had cough at baseline).

Abbreviatures: Time (index -contact): time difference between visit 1 of the TB index case and visit 1 of the contact, PCA: Principal Component Analysis, IGRA: Interferon-Gamma Release Assay, TB: Tuberculosis.

### **Supplementary Table 6** . Characteristics of TB contacts with positive IGRA result at baseline and did not start treatment or received <30 days and TB index cases by TB progression status.

| Characteristics | n/N | No progression to TB (n=270) | Progression to TB (n=15) | p-value |
| --- | --- | --- | --- | --- |
| TB contacts |  |  |  |  |
| Age – median (IQR) | 285/285 | 38.8 (24.1-68.6) | 26.6 (20.2-61.7) | 0.412 |
| Female– no. (%) | 285/285 | 183 (67.8) | 10 (66.7) | 1 |
| Race/Ethnicity – no. (%) | 284/285 |  |  | 0.417 |
| White |  | 32 (11.9) | 4 (26.7) |  |
| Black |  | 98 (36.4) | 4 (26.7) |  |
| Asian |  | 1 (0.4) | 0 (0) |  |
| Pardo |  | 135 (50.2) | 7 (46.7) |  |
| Indigenous |  | 3 (1.1) | 0 (0) |  |
| BMI – median (IQR) | 284/285 | 25.3 (22.1-36.6) | 24.2 (20.4-28.1) | **0.04** |
| Literate– no. (%) | 284/285 | 255 (94.8) | 14 (93.3) | 0.566 |
| Income – no. (%) | 277/285 |  |  | 0.240 |
| Without income |  | 52 (19.8) | 5 (35.7) |  |
| Equal or less than a minimum wage |  | 138 (52.5) | 6 (42.9) |  |
| More than a minimum wage |  | 73 (27.8) | 3 (21.4) |  |
| BCG scar – no. (%) | 284/285 | 238 (88.5) | 14 (93.3) | 0.563 |
| HIV infection – no. (%) | 285/285 | 2 (0.7) | 0 (0) | NA |
| ART (if HIV+) – no. (%) | 2/285 | 1 (50) | 0 (0) | NA |
| Smoking – no. (%) | 284/285 |  |  | 0.165 |
| Current |  | 45 (16.7) | 1 (6.7) |  |
| Former |  | 39 (14.5) | 1 (6.7) |  |
| Never |  | 185 (68.8) | 13 (86.7) |  |
| Passive smoking – no. (%) | 282/285 | 112 (41.9) | 7 (46.7) | 0.797 |
| Alcohol consumption – no. (%) | 284/285 |  |  | 0.113 |
| Current |  | 107 (39.8) | 4 (26.7) |  |
| Former |  | 50 (18.6) | 1 (6.7) |  |
| Never |  | 112 (41.6) | 10 (66.7) |  |
| drug use – no. (%) | 284/285 |  |  | NA |
| Current |  | 3 (1.1) | 0 (0) |  |
| Former |  | 15 (5.6) | 0 (0) |  |
| Never |  | 251 (93.3) | 15 (100) |  |
| Diabetes– no. (%) | 281/285 | 14 (5.3) | 0 (0) | NA |
| Hypertension– no. (%) | 281/285 | 40 (15) | 2 (13.3) | 0.857 |
| Time _index-contact_ (weeks) – median (IQR) | 281/285 | 17 (7-188) | 15 (4-126) | 0.553 |
| TB index cases |  |  |  |  |
| Age – median (IQR) | 285/285 | 33 (22-64) | 30 (20-64) | 0.859 |
| Female– no. (%) | 285/285 | 176 (65.2) | 9 (60) | 0.782 |
| Multidrug resistance – no. (%) | 284/285 | 3 (1.1) | 0 (0) | NA |
| HbA1c (%)-median (IQR) | 283/285 | 5.8 (5.5-12.1) | 5.8 (5.5-13.8) | 0.499 |
| Enrollment Cough – no. (%) | 285/285 | 260 (96.3) | 15 (100) | 0.448 |
| Chest X-ray– no. (%) | 276/285 | 189 (72.4) | 11 (73.3) | 1 |
| Smoking – no. (%) | 285/285 |  |  | 0.211 |
| Current |  | 84 (31.1) | 3 (20) |  |
| Former |  | 68 (25.2) | 3 (20) |  |
| Never |  | 118 (43.7) | 9 (60) |  |
| Passive smoking – no. (%) | 284/285 | 132 (49.1) | 11 (73.3) | 0.109 |
| Alcohol consumption – no. (%) | 285/285 |  |  | 0.456 |
| Current |  | 124 (45.9) | 6 (40) |  |
| Former |  | 83 (30.7) | 4 (26.7) |  |
| Never |  | 63 (23.3) | 5 (33.3) |  |
| drug use – no. (%) | 285/285 |  |  | 0.188 |
| Current |  | 43 (15.9) | 1 (6.7) |  |
| Former |  | 40 (14.8) | 1 (6.7) |  |
| Never |  | 187 (69.3) | 13 (86.7) |  |
| Positive sputum smear – no. (%) | 271/285 | 226 (88.3) | 13 (86.7) | 0.693 |

**Table note.** Data represent no. (%), except for age and BMI, which is presented as median and interquartile range (IQR). Continuous variables were compared using the Mann Whitney U test and categorical variables were using the Pearson’s chi-square test. Bold values represent statistically significant.

Abbreviations: TB: tuberculosis, BMI: Body Mass Index, ART: antiretroviral therapy, IGRA; Interferon- Gamma Release Assay, TPT: Tuberculosis preventive treatment.

### **Supplementary Table 7.** Characteristics of TB contacts with positive IGRA result at baseline and did not start treatment or received <30 days and TB cases included and excluded in each Dimension Reduction stage using empirical review and LASSO regression.

| **Characteristics** | | **Dimensional Reduction** | |
| --- | --- | --- | --- |
|  |  | **Empirical Review** | **LASSO** |
| **TB contact** | Age | Included | **Excluded** |
|  | Sex | Included | **Excluded** |
|  | Race/Ethnicity | Included | **Excluded** |
|  | Body Mass Index | Included | Included |
|  | Literate | Included | **Excluded** |
|  | Income | Included | **Excluded** |
|  | BCG scar | Included | **Excluded** |
|  | HIV infection | **Excluded** |  |
|  | Antiretroviral therapy (if HIV+) | **Excluded** |  |
|  | Smoking | Included | **Excluded** |
|  | Passive smoking | Included | **Excluded** |
|  | Alcohol consumption | Included | **Excluded** |
|  | CAGE assessment | **Excluded** |  |
|  | drug use | **Excluded** |  |
|  | Diabetes | **Excluded** |  |
|  | Hypertension | **Excluded** |  |
|  | Time (index-contact) | Included | **Excluded** |
| **TB cases** | Age | Included | **Excluded** |
|  | Sex | Included | **Excluded** |
|  | Multidrug resistance | **Excluded** |  |
|  | HbA1c | Included | **Excluded** |
|  | Enrollment Cough | **Excluded** |  |
|  | Cavity on chest X-ray | Included | **Excluded** |
|  | Passive smoking | Included | **Excluded** |
|  | Smoking | Included | **Excluded** |
|  | Alcohol consumption | Included | **Excluded** |
|  | Drug use | Included | **Excluded** |
|  | Positive sputum smear | Included | **Excluded** |

**Table note.** Only variables included in the previous stage were entered into the next stage.

*Eight variables were excluded: HIV infection and antiretroviral therapy (no progressors to TB were HIV+), CAGE assessment ("alcohol consumption" variable was similar), drug use (no progressors to TB used drugs), hypertension and diabetes (were self-reported variables), multidrug resistance (there was no TB index of progressors to TB with multidrug resistance) and cough at baseline (all TB index had cough at baseline).

Abbreviations: Time (index -contact): time difference between visit 1 of the TB index case and visit 1 of the contact, IGRA: Interferon-Gamma Release Assay, TB: Tuberculosis.

### **Supplementary Table 8**. Characteristics of TB contacts with positive IGRA result at baseline and did not start treatment or received <30 days and TB cases included and excluded in each Dimension Reduction stage using empirical review, PCA and LASSO regression.

| **Characteristics** | | **Dimensional Reduction** | | |
| --- | --- | --- | --- | --- |
|  |  | **Empirical Review** | **PCA** | **LASSO** |
| **TB contact** | Age | Included | Included | **Excluded** |
|  | Sex | Included | **Excluded** |  |
|  | Race/Ethnicity | Included | **Excluded** |  |
|  | Body Mass Index | Included | Included | Included |
|  | Literate | Included | **Excluded** |  |
|  | Income | Included | **Excluded** |  |
|  | BCG scar | Included | Included | **Excluded** |
|  | HIV infection | **Excluded** |  |  |
|  | Antiretroviral therapy (if HIV+) | **Excluded** |  |  |
|  | Smoking | Included | Included | **Excluded** |
|  | Passive smoking | Included | Included | **Excluded** |
|  | Alcohol consumption | Included | Included | **Excluded** |
|  | CAGE assessment | **Excluded** |  |  |
|  | drug use | **Excluded** |  |  |
|  | Diabetes | **Excluded** |  |  |
|  | Hypertension | **Excluded** |  |  |
|  | Time (index-contact) | Included | Included | **Excluded** |
| **TB cases** | Age | Included | Included | **Excluded** |
|  | Sex | Included | **Excluded** |  |
|  | Multidrug resistance | **Excluded** |  |  |
|  | HbA1c | Included | Included | **Excluded** |
|  | Enrollment Cough | **Excluded** |  |  |
|  | Cavity on chest X-ray | Included | **Excluded** |  |
|  | Passive smoking | Included | Included | Included |
|  | Smoking | Included | **Excluded** |  |
|  | Alcohol consumption | Included | Included | **Excluded** |
|  | Drug use | Included | Included | **Excluded** |
|  | Positive sputum smear | Included | Included | **Excluded** |

**Table note.** Only variables included in the previous stage were entered into the next stage.

*Eight variables were excluded: HIV infection and antiretroviral therapy (no progressors to TB were HIV+), CAGE assessment ("alcohol consumption" variable was similar), drug use (no progressors to TB used drugs), hypertension and diabetes (were self-reported variables), multidrug resistance (there was no TB index of progressors to TB with multidrug resistance) and cough at baseline (all TB index had cough at baseline).

Abbreviations: Time (index -contact): time difference between visit 1 of the TB index case and visit 1 of the contact, PCA: Principal Components Analysis, IGRA: Interferon-Gamma Release Assay, TB: Tuberculosis.

### **Supplementary Table 9**. Mixed-effects Cox proportional hazard model for progression to TB among all close contacts enrolled in the study

| **Predictors** | **adjusted Hazard Ratio** | **95% (CI)** | **p-value** |
| --- | --- | --- | --- |
| **Time (index -contact)** | 1.00 | 0.99 – 1.00 | 0.241 |
| ***Tuberculosis preventive treatment*** |  |  |  |
| Never started | 13.49 | 1.78 – 102.10 | **0.012** |
| Incomplete | 6.90 | 0.72 – 66.31 | 0.095 |
| Completed | 1.50 | 0.12 – 18.78 | 0.751 |
| Not recommended | reference | | |
| ***Positive IGRA result at baseline*** | 1.64 | 0.48 – 5.66 | 0.430 |

Optimism-corrected Area under the curve of this model: 0.82 (95% CI: 0.72-0.90) obtained after 200 bootstrap replications

Dimension reduction (empirical review, PCA and Lasso regression) was used to select the variables included in this model.

Abbreviations: Time (index -contact): time difference between visit 1 of the TB index case and visit 1 of the contact, IGRA: Interferon-Gamma Release Assay, TB: Tuberculosis, CI: Confidence Interval, PCA: Principal Component Analysis,

### **Supplementary Table 10**. Mixed-effects Cox proportional hazard model for progression to TB among contacts with positive IGRA result at baseline enrolled in the study

| **Predictors** | **adjusted Hazard Ratio** | **95% (CI)** | **p-value** |
| --- | --- | --- | --- |
| ***Tuberculosis preventive treatment*** |  |  |  |
| Never started | 13.12 | 1.72 – 99.75 | **0.013** |
| Incomplete | 2.30 | 0.14 – 36.75 | 0.556 |
| Not recommended | reference | | |

Optimism-corrected Area under the curve of this model: 0.74 (95% CI: 0.66-0.80) obtained after 200 bootstrap replications

Dimension reduction (empirical review, PCA and Lasso regression) was used to select the variables included in this model.

Abbreviations: TB: Tuberculosis, CI: Confidence Interval, PCA: Principal Component Analysis,

### **Supplementary Table 11**. Mixed-effects Cox proportional hazard model for progression to TB among close contacts with a positive IGRA result at baseline who did not start treatment or received <30 days enrolled in the study.

| **Predictors** | **adjusted Hazard Ratio** | **95% (CI)** | **p-value** |
| --- | --- | --- | --- |
| ***BMI (kg/m2) (TB contact)*** | 0.88 | 0.78 – 0.98 | **0.017** |
| ***Secondary smoking (TB index case)*** | 0.38 | 0.12 – 1.20 | 0.101 |

Optimism-corrected Area under the curve of this model: 0.72 (95% CI: 0.61-0.84) obtained after 200 bootstrap replications

Dimension reduction (empirical review, PCA and Lasso regression) was used to select the variables included in this model.

Abbreviations: TB: Tuberculosis, BMI: Body mass index, CI: Confidence Interval, PCA: Principal Component Analysis,

### **Supplementary Table 12.** Mixed-effects Cox proportional hazard model for progression to TB among all close contacts enrolled in the study with imputed data.

| **Predictors** | **adjusted Hazard Ratio** | **95% (CI)** | **p-value** |
| --- | --- | --- | --- |
| ***Tuberculosis preventive treatment*** |  |  |  |
| Never started | 15.77 | 2.10 – 118.46 | **0.007** |
| Incomplete | 6.78 | 0.71 – 65.22 | 0.097 |
| Completed | 1.48 | 0.13 – 17.43 | 0.756 |
| Not recommended | reference | | |
| ***Positive IGRA result at baseline*** | 1.62 | 0.54 – 4.81 | 0.387 |

Optimism-corrected Area under the curve of this model: 0.80 (95% CI: 0.71-0.88) obtained after 200 bootstrap replications.

Dimension reduction (empirical review and Lasso regression) was used to select the variables included in this model.

Abbreviations: IGRA: Interferon-Gamma Release Assay, TB: Tuberculosis, CI: Confidence Interval

### **Supplementary Table 13.** Mixed-effects Cox proportional hazard model for progression to TB among contacts with positive IGRA result at baseline enrolled in the study with imputed data

| **Predictors** | **adjusted Hazard Ratio** | **95% (CI)** | **p-value** |
| --- | --- | --- | --- |
| ***Tuberculosis preventive treatment*** |  |  |  |
| Never started | 17.25 | 2.27 – 131.26 | **0.006** |
| Incomplete | 5.00 | 0.45 – 55.28 | 0.189 |
| Not recommended | reference | | |
| ***Age (years) (TB index case)*** | 1.01 | 0.98 – 1.04 | 0.495 |
| ***Secondary smoking (TB index case)*** | 0.47 | 0.17 – 1.34 | 0.159 |
| ***Use illicit drug (TB index case)*** | 0.39 | 0.05 – 3.05 | 0.371 |

Optimism-corrected Area under the curve of this model: 0.79 (95% CI: 0.70-0.87) obtained after 200 bootstrap replications

Dimension reduction (empirical review and Lasso regression) was used to select the variables included in this model.

Abbreviations: TB: Tuberculosis, CI: Confidence Interval

### **Supplementary Table 14.** Mixed-effects Cox proportional hazard model for progression to TB among close contacts with a positive IGRA result at baseline who did not start treatment or received <30 days enrolled in the study with imputed data.

| **Predictors** | **adjusted Hazard Ratio** | **95% (CI)** | **p-value** |
| --- | --- | --- | --- |
| ***BMI (kg/m^2^) (TB contact)*** | 0.88 | 0.79 – 0.98 | **0.022** |
| ***Black/Pardo race (TB contact)*** | 0.38 | 0.12 – 1.23 | 0.106 |
| ***Secondary smoking (TB index case)*** | 0.32 | 0.10 – 1.02 | 0.053 |

Optimism-corrected Area under the curve of this model: 0.75 (95% CI: 0.63-0.85) obtained after 200 bootstrap replications

Dimension reduction (empirical review and Lasso regression) was used to select the variables included in this model.

Abbreviations: TB: Tuberculosis, BMI: Body mass index, CI: Confidence Interval
